# Sex influences the effects of APOE genotype and Alzheimer’s diagnosis on neuropathology and memory

**DOI:** 10.1101/2020.06.25.20139980

**Authors:** Paula Duarte-Guterman, Arianne Y. Albert, Cindy K. Barha, Liisa A.M. Galea on behalf of the Alzheimer’s Disease Neuroimaging Initiative

## Abstract

Alzheimer’s disease (AD) is characterised by severe cognitive decline and pathological changes in the brain (brain atrophy, hyperphosphorylation of tau, and deposition of toxic amyloid-beta protein). Females have greater neuropathology (AD biomarkers and brain atrophy rates) and cognitive decline than males, however these effects can depend on diagnosis (amnestic mild cognitive impairment (aMCI) or AD) and APOE genotype (presence of ε4 alleles). Using the ADNI database (N=630 females, N=830 males), we analysed the effect of sex, APOE genotype (non-carriers or carriers of APOEε4 alleles), and diagnosis (cognitively normal (CN), early aMCI (EMCI), late aMCI (LMCI), probable AD) on cognition (memory and executive function), hippocampal volume, and AD biomarkers (CSF levels of amyloid beta, tau and ptau). Regardless of APOE genotype, memory scores were higher in CN, EMCI, and LMCI females compared to males but this sex difference was absent in probable AD, which may suggest a delay in the onset of cognitive decline or diagnosis and/or a faster trajectory of cognitive decline in females. We found that, regardless of diagnosis, CSF tau-pathology was disproportionately elevated in female carriers of APOEε4 alleles compared to males. In contrast, male carriers of APOEε4 alleles had reduced levels of CSF amyloid beta compared to females, irrespective of diagnosis. We also detected sex differences in hippocampal volume but the direction was dependent on the method of correction. Altogether results suggest that across diagnosis females show greater memory decline compared to males and APOE genotype affects AD neuropathology differently in males and females which may influence sex differences in incidence and progression of aMCI and AD.

## 1. Introduction

Alzheimer’s disease (AD) is characterized by severe cognitive decline and neuropathological markers such as brain atrophy, hyperphosphorylation of tau, and deposition of toxic amyloid-beta (Aβ) protein in the brain (Alzheimer’s Association, 2017). The hippocampus is one of the first brain areas to show atrophy with AD (Jack et al., 2000; Kidron et al., 1997) and hippocampal atrophy correlates with cognitive decline (Petersen et al., 2000) and AD pathology (neurofibrillary tangles; Jack et al., 2002). Possession of one or two APOEε4 alleles, the strongest genetic risk factor for sporadic AD, and female sex are important non-modifiable risk factors for AD (Riedel et al., 2016). Studies indicate that females with AD show greater signs of neuropathology (tau levels), rates of brain atrophy, including in the hippocampus, and cognitive decline than males, which may be exacerbated by APOE genotype (e.g., Ardekani et al., 2016; Buckley et al., 2018; Cavedo et al., 2018; Hohman et al., 2018). One allele of APOEε4 increases the risk of AD in females relative to males between 65 and 75 years, indicating that the APOE genotype affects males and females differently (Neu et al., 2017).

Previous studies have examined the interaction of APOE genotype and sex on AD biomarkers (tau and Aβ), hippocampal atrophy, and cognitive function, but findings are not consistent (Altmann et al., 2014; Buckley et al., 2018; Damoiseaux et al., 2012; Holland et al., 2013; Liu et al., 2019; Sampedro et al., 2015; Sohn et al., 2018; Wang et al., 2019), likely due to the diagnosis groups included and whether studies use longitudinal or baseline comparisons. Using the Alzheimer’s Disease Neuroimaging Initiative (ADNI) database, previous studies found a stronger association between APOEε4 and CSF tau levels in females compared to males at baseline in cognitively normal (CN; Damoiseaux et al., 2012; but see Sampedro et al., 2015) and amnestic mild cognitive impairment (aMCI) individuals (Altmann et al., 2014; Liu et al., 2019) but no studies to our knowledge have investigated the interactions between sex and APOE genotype in individuals with AD. Females with APOEε4 alleles have steeper hippocampal volume reductions with age in CN individuals (Holland et al., 2013) and show greater cognitive decline in aMCI individuals (Sohn et al., 2018; Wang et al., 2019) compared to males but these findings are not consistent (Buckley et al., 2018). Although there are several ADNI studies that have examined sex by APOE genotype interactions on AD biomarkers, most of these studies included only one diagnosis group, with either only CN (Buckley et al., 2018, 2019a) or aMCI groups (Liu et al., 2019; Sohn et al., 2018) or analysed the groups (CN, aMCI, AD) separately (Sundermann et al., 2018). It is important to include all three diagnosis groups CN, aMCI and AD to understand how sex may interact with APOE genotype to influence AD biomarkers across diagnosis. In order to improve diagnosis and treatment, it is important to understand why females are at a higher lifetime risk and have a higher burden of the disease than males and whether this is across diagnosis and APOE genotype for a number of AD biomarkers.

Sex differences in memory depend on memory domain, females perform better in episodic memory tasks related to verbal memory, whereas males perform better in visuospatial-related tasks (meta-analysis by Asperholm et al., 2019). Furthermore, there are sex differences in some forms of executive function depending on the type of executive function being examined, with a male advantage for working memory and a female advantage on response inhibition in cognitively healthy adults (Gaillard et al., 2020). In tasks that involve the integrity of the hippocampus, males typically outperform females in both humans and rodents (Yagi and Galea, 2019). In addition, while sex differences in hippocampal volume have been detected, with females having a smaller volume compared to males, this depends on whether the data are corrected for individual differences in brain volume and the method of correction employed (meta-analysis by Tan et al., 2016). Understanding basic sex differences in hippocampal function and structure is important to determine how they can contribute to sex differences in vulnerability and progression of neurodegenerative diseases in which the integrity of the hippocampus is affected such as in AD.

The objective of this study was to examine sex differences in AD biomarkers (CSF Aβ, tau, and phosphorylated tau), volume of the hippocampus (using two correction factors), memory, and executive function, and how these may be affected by APOE genotype (non-carriers or carriers of APOEε4 alleles) and diagnosis status (CN, aMCI, probable AD). We hypothesized that females will be more affected by APOE genotype and diagnosis and this will be reflected in greater AD pathology, smaller hippocampus volume, and lower cognitive scores than males with probable AD and this is exacerbated in female carriers of APOEε4 alleles.

## 2. Materials and methods

### 2.1 ADNI database

Data used in the preparation of this article were obtained from the Alzheimer’s Disease Neuroimaging Initiative (ADNI) database (adni.loni.usc.edu). The ADNI was launched in 2003 as a public-private partnership, led by Principal Investigator Michael W. Weiner, MD. The primary goal of ADNI has been to test whether serial magnetic resonance imaging (MRI), positron emission tomography (PET), other biological markers, and clinical and neuropsychological assessment can be combined to measure the progression of mild cognitive impairment (MCI) and early Alzheimer’s disease (AD). For up-to-date information, see www.adni-info.org. Data used in this article were downloaded on or before Jan 16, 2019. Participants (between the ages of 55 and 90) were recruited across the United States and Canada and agreed to complete a variety of imaging and clinical assessments at baseline and repeated at specific intervals (Petersen et al., 2010). The study design and schedule is available online: (http://adni.loni.usc.edu/study-design/). Inclusion and exclusion criteria are detailed online (http://adni.loni.usc.edu/methods/documents/). Briefly, CN participants had normal memory function (measured by education-adjusted scores on the Wechsler Memory Scale Logical Memory II) and a Clinical Dementia Rating (CDR) of 0. Amnestic late MCI (LMCI) participants had objective memory loss (from Wechsler Memory Scale Logical Memory II scores), a CDR of 0.5, preserved daily activities, and absence of dementia. Early MCI (EMCI) participants had a milder episodic memory impairment compared to LMCI (Wechsler Memory Scale Logical Memory II scores ∼0.5 and 1.5 SD below the mean of CN participants). Participants that met NINCDS/ADRDA Alzheimer’s Criteria and a CDR of 0.5 or 1.0 were categorised as probable AD.

### 2.2 CSF biomarkers, hippocampal volume, memory, and executive function

We included all participants who had the following baseline data: diagnosis in the ADNI database, cerebrospinal fluid (CSF) levels for Aβ, tau and phosphorylated tau (ptau), had a brain MRI scan, and underwent a battery of neuropsychological tests (total n = 1,460, n= 630 females, n=830 males; Table 1). Data included in our analyses were: demographics (age, years of education, and ethnicity), baseline diagnosis (cognitively normal, CN; EMCI; LMCI; or probable AD), number of APOE ε4 alleles (0, 1 or 2), CSF Aβ (pg/ml), CSF tau (pg/ml), and CSF p-tau (pg/ml), hippocampal volume (mm^3^), ADNI executive function z-scores, and ADNI memory z-scores (using data from the ADNI neuropsychological battery and validated in Crane et al., 2012; Gibbons et al., 2012). The executive function score included WAIS-R Digit Symbol Substitution, Digit Span Backwards, Trails A and B, Category Fluency, and Clock Drawing (Gibbons et al., 2012). The composite memory score included Rey Auditory Verbal Learning Test, AD Assessment Schedule - Cognition, Mini-Mental State Examination, and Logical Memory data (Crane et al., 2012). APOE genotype was determined from EDTA blood samples collected at baseline (for a detailed protocol see: Shaw et al., 2009). Tau and Aβ levels were determined in CSF collected in the morning after an overnight fast and using the micro-bead-based multiplex immunoassay, the INNO-BIA AlzBio3 RUO test (Fujirebio, Ghent, Belgium) on the Luminex platform (for details see: Shaw et al., 2009). MRI scans were obtained according to standardised protocol (http://adni.loni.usc.edu/methods/mri-analysis/mri-acquisition/). Hippocampal volume data were analysed using FreeSurfer (https://surfer.nmr.mgh.harvard.edu) version 4.3 for ADNI 1 and version 5.1 for ADNI GO and 2 at the University of California – San Francisco (http://adni.loni.usc.edu/methods/). Sex differences in hippocampal volume are influenced by controlling factors such as intracranial volume (Lotze et al., 2019; Tan et al., 2016). In the present study, we corrected hippocampus volume using two different methods. The first method (regression method) used the method in Mormino et al. (2014) and Jack et al. (2012) using the residuals of the linear regression between hippocampal volume and total intracranial volume. We also compared another widely used volume correction method (Sohn et al., 2018; Sundermann et al., 2018; meta-analysis by Tan et al., 2016) by dividing hippocampal volume with intracranial volume to directly compare the different correction methods.

**Table 1.**
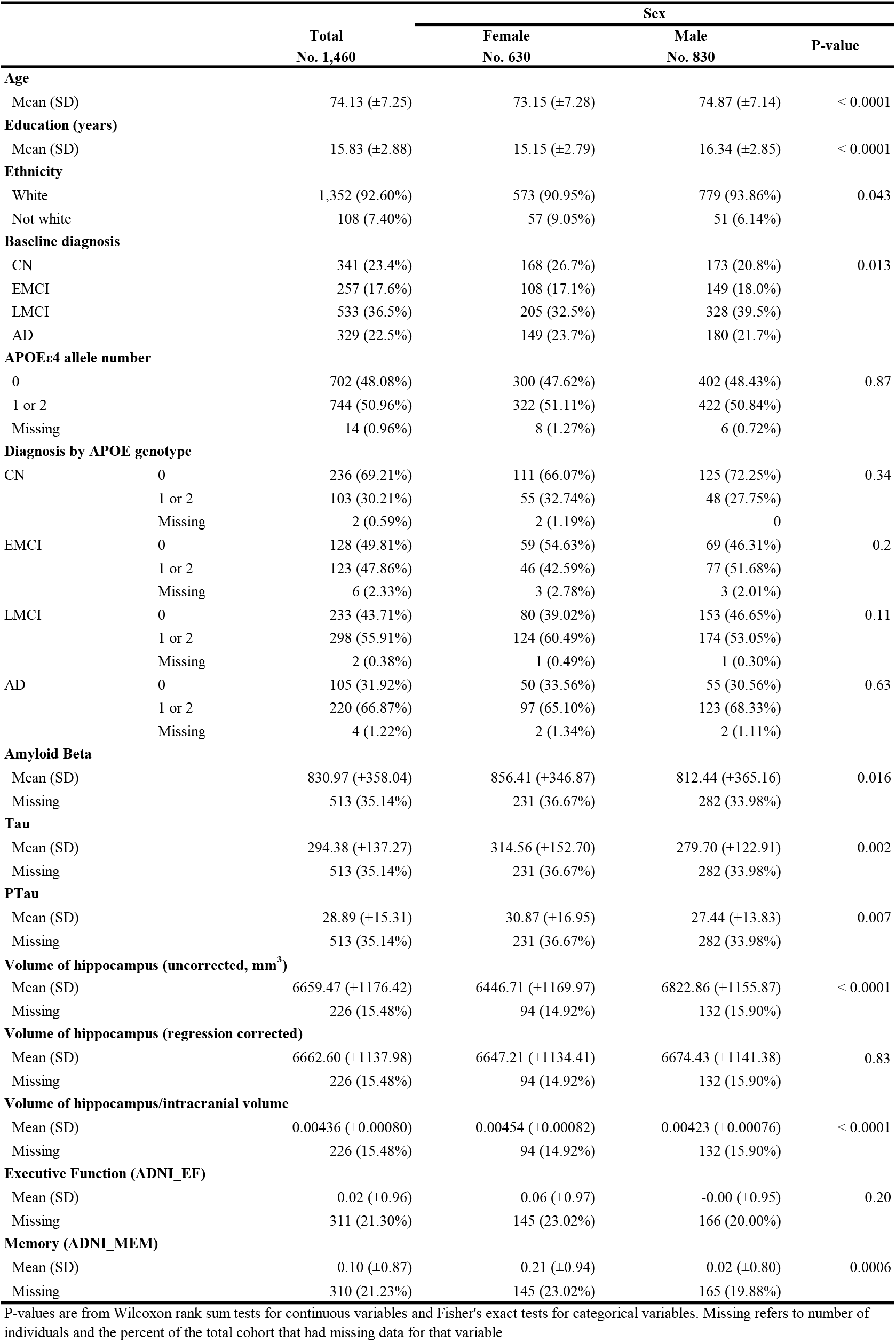
Demographic and clinical information for all participants and subdivided by sex. We collapsed APOE genotype into two groups: (1) participants with no ε4 risk alleles (-/-) and (2) participants carrying any ε4 alleles (homozygous ε4/ε4 and heterozygous ε4/-). Biomarkers for AD are from cerebrospinal fluid. Volume of the hippocampus was corrected using a regression method or using a ratio with intracranial volume (see methods). CN, cognitively normal; EMCI, early mild cognitive impairment; LMCI, late mild cognitive impairment; AD, Alzheimer’s disease.

|  |  | Sex |  |  |  |
| --- | --- | --- | --- | --- | --- |
|  |  | Total | Female | Male | P-value |
|  |  | No. 1,460 | No. 630 | No. 830 |  |
| Age |  |  |  |  |  |
| Mean (SD) |  | 74.13 (±7.25) | 73.15 (±7.28) | 74.87 (±7.14) | < 0.0001 |
| Education (years) |  |  |  |  |  |
| Mean (SD) |  | 15.83 (±2.88) | 15.15 (±2.79) | 16.34 (±2.85) | < 0.0001 |
| Ethnicity |  |  |  |  |  |
| White |  | 1,352 (92.60%) | 573 (90.95%) | 779 (93.86%) | 0.043 |
| Not white |  | 108 (7.40%) | 57 (9.05%) | 51 (6.14%) |  |
| Baseline diagnosis |  |  |  |  |  |
| CN |  | 341 (23.4%) | 168 (26.7%) | 173 (20.8%) | 0.013 |
| EMCI |  | 257 (17.6%) | 108 (17.1%) | 149 (18.0%) |  |
| LMCI |  | 533 (36.5%) | 205 (32.5%) | 328 (39.5%) |  |
| AD |  | 329 (22.5%) | 149 (23.7%) | 180 (21.7%) |  |
| APOEε4 allele number |  |  |  |  |  |
| 0 |  | 702 (48.08%) | 300 (47.62%) | 402 (48.43%) | 0.87 |
| 1 or 2 |  | 744 (50.96%) | 322 (51.11%) | 422 (50.84%) |  |
| Missing |  | 14 (0.96%) | 8 (1.27%) | 6 (0.72%) |  |
| Diagnosis by APOE genotype |  |  |  |  |  |
| CN | 0 | 236 (69.21%) | 111 (66.07%) | 125 (72.25%) | 0.34 |
|  | 1 or 2 | 103 (30.21%) | 55 (32.74%) | 48 (27.75%) |  |
|  | Missing | 2 (0.59%) | 2 (1.19%) | 0 |  |
| EMCI | 0 | 128 (49.81%) | 59 (54.63%) | 69 (46.31%) | 0.2 |
|  | 1 or 2 | 123 (47.86%) | 46 (42.59%) | 77 (51.68%) |  |
|  | Missing | 6 (2.33%) | 3 (2.78%) | 3 (2.01%) |  |
| LMCI | 0 | 233 (43.71%) | 80 (39.02%) | 153 (46.65%) | 0.11 |
|  | 1 or 2 | 298 (55.91%) | 124 (60.49%) | 174 (53.05%) |  |
|  | Missing | 2 (0.38%) | 1 (0.49%) | 1 (0.30%) |  |
| AD | 0 | 105 (31.92%) | 50 (33.56%) | 55 (30.56%) | 0.63 |
|  | 1 or 2 | 220 (66.87%) | 97 (65.10%) | 123 (68.33%) |  |
|  | Missing | 4 (1.22%) | 2 (1.34%) | 2 (1.11%) |  |
| Amyloid Beta |  |  |  |  |  |
| Mean (SD) |  | 830.97 (±358.04) | 856.41 (±346.87) | 812.44 (±365.16) | 0.016 |
| Missing |  | 513 (35.14%) | 231 (36.67%) | 282 (33.98%) |  |
| Tau |  |  |  |  |  |
| Mean (SD) |  | 294.38 (±137.27) | 314.56 (±152.70) | 279.70 (±122.91) | 0.002 |
| Missing |  | 513 (35.14%) | 231 (36.67%) | 282 (33.98%) |  |
| PTau |  |  |  |  |  |
| Mean (SD) |  | 28.89 (±15.31) | 30.87 (±16.95) | 27.44 (±13.83) | 0.007 |
| Missing |  | 513 (35.14%) | 231 (36.67%) | 282 (33.98%) |  |
| Volume of hippocampus (uncorrected, mm<sup>3</sup>) |  |  |  |  |  |
| Mean (SD) |  | 6659.47 (±1176.42) | 6446.71 (±1169.97) | 6822.86 (±1155.87) | < 0.0001 |
| Missing |  | 226 (15.48%) | 94 (14.92%) | 132 (15.90%) |  |
| Volume of hippocampus (regression corrected) |  |  |  |  |  |
| Mean (SD) |  | 6662.60 (±1137.98) | 6647.21 (±1134.41) | 6674.43 (±1141.38) | 0.83 |
| Missing |  | 226 (15.48%) | 94 (14.92%) | 132 (15.90%) |  |
| Volume of hippocampus/intracranial volume |  |  |  |  |  |
| Mean (SD) |  | 0.00436 (±0.00080) | 0.00454 (±0.00082) | 0.00423 (±0.00076) | < 0.0001 |
| Missing |  | 226 (15.48%) | 94 (14.92%) | 132 (15.90%) |  |
| Executive Function (ADNI_EF) |  |  |  |  |  |
| Mean (SD) |  | 0.02 (±0.96) | 0.06 (±0.97) | -0.00 (±0.95) | 0.20 |
| Missing |  | 311 (21.30%) | 145 (23.02%) | 166 (20.00%) |  |
| Memory (ADNI_MEM) |  |  |  |  |  |
| Mean (SD) |  | 0.10 (±0.87) | 0.21 (±0.94) | 0.02 (±0.80) | 0.0006 |
| Missing |  | 310 (21.23%) | 145 (23.02%) | 165 (19.88%) |  |

P-values are from Wilcoxon rank sum tests for continuous variables and Fisher's exact tests for categorical variables. Missing refers to number of individuals and the percent of the total cohort that had missing data for that variable

### 2.3 Statistical Methods

We compared all available data for each study variable between the sexes using the Wilcoxon rank sum test for continuous variables and Fisher’s exact test for categorical variables. We used general linear models to determine the relationships between sex, APOE genotype (non-carriers or carriers of APOEε4 alleles), and diagnosis status (CN, EMCI, LMCI, and probable AD) as predictor variables, and AD biomarkers, corrected hippocampal volume, and cognitive ability, as dependent variables. All models included age and years of education as a covariate. All models initially included the three-way interaction between sex, APOE genotype, and diagnosis; if this interaction was not significant, it was removed from the model to estimate the two way interactions (sex and APOE genotype, sex and diagnosis, APOE genotype and diagnosis). If no two-way interactions were significant, these were removed from the model to estimate the main effects of sex, APOE genotype, and diagnosis. Significance was based on the likelihood ratio test, and all p-values were corrected for multiple testing using the Benjamini-Hochberg false discovery rate method with the family-wise error rate set to 0.05 (Benjamini and Hochberg, 1995). In total, seven p-values per dependent variable were included in each set of models (interaction terms and main effects of sex, APOE, and diagnosis) resulting in 49 p-values corrected (seven dependent variables; Tables 1 and 2). Significant interaction terms were followed up using pairwise simple-effects tests with Benjamini-Hochberg p-value correction. We calculated Pearson’s correlation coefficients between CSF levels of tau and Aβ by sex and APOE genotype separately. We report significance differences (adjusted p≤0.05). All regression analyses were carried out in R v3.5.1 (R Core Team, 2018).

**Table 2.**
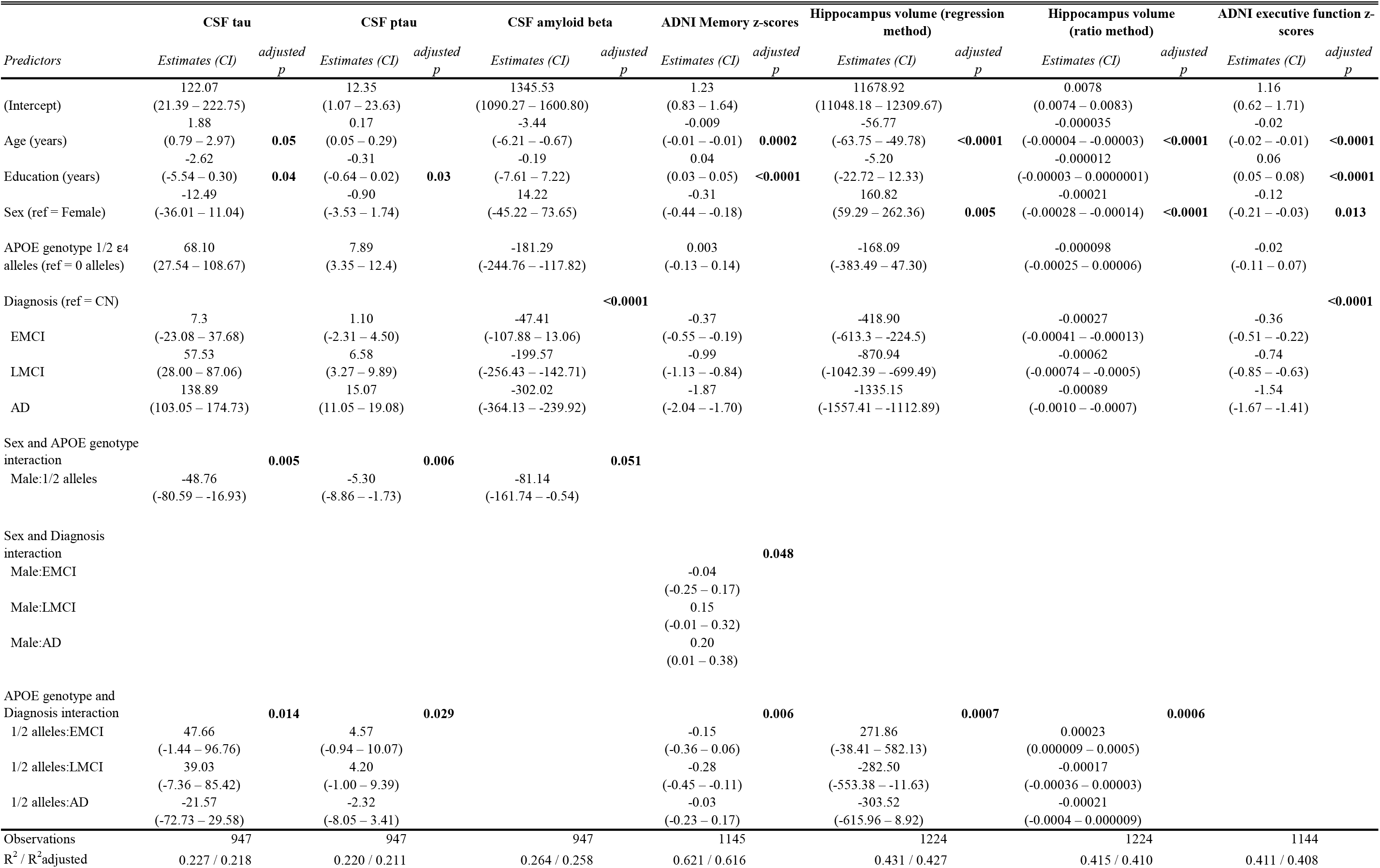
Linear regression results for models with sex, baseline diagnosis (CN, cognitively normal; EMCI, early mild cognitive impairment; LMCI, late mild cognitive impairment; AD, Alzheimer’s disease), and APOE4 genotype (non-carriers or carriers of APOEε4 alleles). P-values are for overall tests and are FDR-adjusted. The three-way interactions were not significant for any of the endpoints.

## 3. Results

### 3.1 Demographic information

Table 1 gives a summary of the variables in the data set (N=1460). Overall, females were significantly younger and had fewer years of education than males (p’s<0.0001) and hence age and education were used as covariates in all analyses. There were more white males than white females in our sample and more non-white females compared to non-white males (p<0.05). In terms of APOE genotype, there were no sex differences in distribution of APOE genotype with 40% females and 38.8% males possessing one allele of APOEε4, and 11% females and 12% of males possessing two alleles of APOEε4. The proportion of participants in each of the diagnosis categories was significantly different for females and males (p<0.05). More females were cognitively normal than males (26.7% compared to 20.8%, unadjusted p=0.01) but there were no sex differences in baseline diagnosis of probable AD (females: 23.7% compared to males: 21.7%, unadjusted p=0.41). However, there were more males with a diagnosis of LMCI (39.5% versus 32.5%, unadjusted p=0.007) but not EMCI (18.0% versus 17.1%, unadjusted p=0.74) compared to females.

We tested whether sex, APOE genotype (non-carriers or carriers of APOEε4 alleles), and diagnosis status (CN, EMCI, LMCI, and probable AD) influenced cognitive ability, corrected hippocampal volume, and CSF biomarkers of AD in one model. However, we did not find significant interactions between these three factors in any of the models (i.e. none of the three-way interactions were significant). We did find significant two-way interactions between sex and APOE genotype, sex and diagnosis, and APOE genotype and diagnosis (Table 2) on these dependent variables, which are discussed in turn below.

### 3.2. Sex and APOE genotype were associated with changes in CSF AD biomarkers

We found significant interactions between sex and APOE genotype for CSF tau, and p-tau (p=0.005 and 0.006, respectively), and a trend for CSF Aβ (p=0.051; Table 2). Tau and p-tau levels were significantly higher in female carriers of APOEε4 alleles compared to male carriers (all p’s<0.0001; Fig. 1 A and B) but no sex differences were detected in non-carriers of APOEε4 alleles (p>0.3). Although CSF tau and p-tau levels were also higher in males with APOEε4 genotype, the difference in levels between carriers and non-carriers was more pronounced in females (standardized regression coefficients for tau and ptau: 0.46 and 0.49 in females and 0.19 and 0.24 in males). For CSF Aβ, post-hoc comparisons showed that female APOEε4 carriers had higher levels compared to males carriers (p=0.04) but there were no sex differences in non-carriers (p=0.6; Fig. 1 C). Levels of CSF Aβ were lower in both sexes in carriers of APOEε4 alleles but the difference between carriers and non-carriers was more pronounced in males than in females, irrespective of diagnosis (standardized regression coefficients for Aβ: −0.58 in females and −0.71 in males). There were no significant sex by diagnosis interactions for AD biomarkers (Table 2).

**Figure 1.**
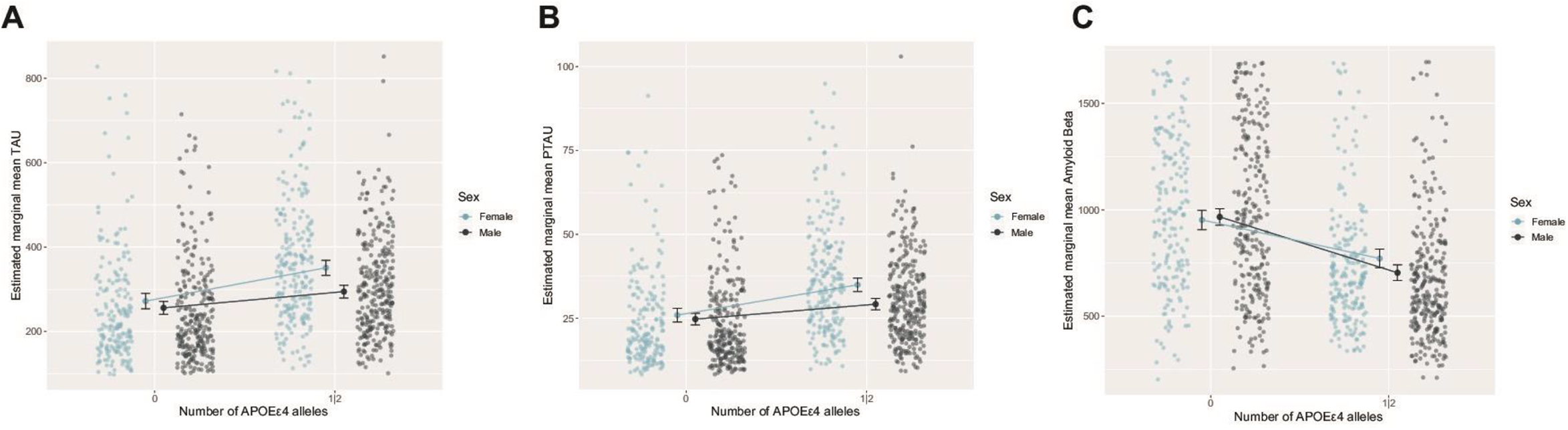
A. Levels of CSF AD biomarkers tau (pg/ml; A), p-tau (pg/ml; B), and amyloid beta (pg/ml; C) in ADNI participants by sex and APOE genotype (absence or presence of APOEε4 alleles).

We next investigated the relationship, using correlations, between CSF tau and Aβ levels in males and females and by APOE genotype (Table 3). We found significant negative correlations between CSF levels of tau and Aβ in male and female non-carriers of APOEε4 alleles (r=-0.262, p<0.0001; r=-0. 195, p=0.008, respectively). However, in carriers of APOEε4 alleles, a significant negative correlation was found only in females (r=-0.219, p=0.001) but not in males (r= −0.046, p=0.44).

**Table 3.** Correlations between CSF levels of tau and amyloid beta, CSF tau and memory scores, and CSF amyloid beta and memory scores, depending by sex and APOE genotype (non carriers versus carriers of ε4 alleles)

| Variables | APOE genotype | Correlation in females | p-value | Correlation in males | p-value |
| --- | --- | --- | --- | --- | --- |
| **CSF tau and Aβ** |  |  |  |  |  |
|  | Non carriers | -0.195 (-0.330 to - 0.053) | **0.008** | -0.262 (-0.372 to -0.145) | **< 0.0001** |
| | Carriers of APOE$\epsilon 4$ alleles | -0.219 (-0.343 to -0.088) | **0.001** | -0.046 (-0.160 to 0.070) | 0.44 |
| **CSF tau and memory z-scores** |  |  |  |  |  |
|  | Non carriers | -0.397 (-0.532 to -0.244) | **< 0.0001** | -0.394 (-0.507 to -0.268) | **< 0.0001** |
| | Carriers of APOE$\epsilon 4$ alleles | -0.242 (-0.387 to -0.086) | **0.003** | -0.232 (-0.355 to -0.100) | **0.0007** |
| **CSF Aβ and memory z-scores** |  |  |  |  |  |
|  | Non carriers | 0.425 (0.275 to 0.555) | **< 0.0001** | 0.305 (0.172 to 0.428) | **< 0.0001** |
| | Carriers of APOE$\epsilon 4$ alleles | 0.407 (0.265 to 0.533) | **< 0.0001** | 0.413 (0.295 to 0.519) | **< 0.0001** |

### 3.3. Sex and diagnosis were associated with changes in memory scores

We found a significant interaction between sex and diagnosis for ADNI memory z-scores (p=0.048; Table 2). Females had significantly higher memory z-scores than males in the CN, EMCI, and LMCI groups (p<0.001, p<0.001, p<0.01, respectively) while no sex differences were detected in probable AD (p=0.1). Thus, severity of diagnosis decreased memory scores to a greater extent in females, such that there was no longer a sex differences in memory scores in individuals with AD (Fig. 2). CSF tau and Aβ were each significantly correlated with memory z-scores in both sexes, regardless of genotype (Table 3). There were no significant sex by APOE genotype interactions for memory scores (Table 2).

**Figure 2.**
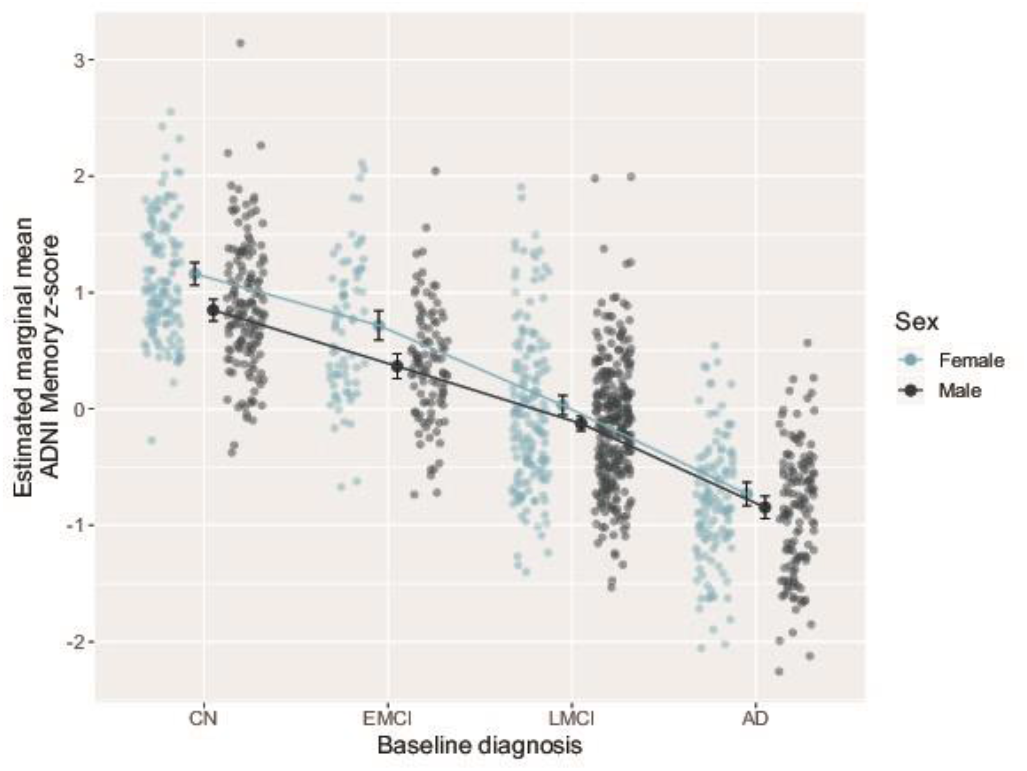
Memory scores in ADNI participants by sex and diagnosis (CN, EMCI, LMCI, AD). CN, cognitively normal; EMCI, early mild cognitive impairment; LMCI, late mild cognitive impairment; AD, Alzheimer’s disease.

### 3.3. Diagnosis and APOE are associated with changes in memory, hippocampal volume, and tau-pathology

Two-way interactions between diagnosis and APOE genotype were detected for memory z-scores (p=0.006), hippocampal volume (the two correction methods; both p’s<0.001), and CSF tau (p=0.014) and ptau levels (p=0.029) but not for executive function scores or CSF Aβ levels. As expected, increasing severity of diagnosis decreases memory z-scores in carriers and non-carriers of APOEε4 alleles (p’s<0.001). However, carrying APOEε4 alleles decreased memory scores in the LCMI group (p<0.0001), but not in the CN, EMCI, and probable AD groups (all p’s>0.2), compared to non-carriers.

For both methods of hippocampal volume calculation, we found that, irrespective of sex, carriers of APOEε4 alleles had a smaller volume compared to non-carriers in LMCI and probable AD individuals (regression method: p<0.0001 and p=0.0004, respectively; ratio method: p=0.0001 and p=0.001, respectively). There were no significant differences in hippocampal volume between carriers and non-carriers of APOEε4 alleles in the EMCI and CN groups (p’s>0.5). In non-carriers, severity of diagnosis was associated with a smaller hippocampal volume, regardless of method of correction (AD< LMCI< EMCI< CN; p’s<0.01). In carriers of APOEε4 alleles, EMCI diagnosis did not significantly affect hippocampal volume compared to CN individuals with both methods of correction (p’s>0.7) while there was a reduction in hippocampal volume in probable AD and LMCI (AD<LMCI<CN; p’s<0.0001). There were no significant sex by diagnosis or sex by APOE genotype interactions for hippocampal volume (Table 2).

CSF tau and ptau levels increased with the severity of diagnosis but this was affected by APOE genotype (p=0.014 and p=0.029, respectively). Carrying APOEε4 alleles resulted in higher levels of CSF tau and ptau in EMCI and LMCI (all p’s<0.0001), but not in CN or probable AD, individuals (p’s>0.17). Regardless of sex, the difference in CSF tau and ptau levels between non-carriers and carriers of APOEε4 alleles was reduced in individuals with probable AD compared to EMCI and LMCI diagnoses.

### 3.5. Sex is associated with hippocampal volume

Males had a larger hippocampus volume by the regression method, but a smaller proportion of the brain was devoted to hippocampus (ratio method) when compared to females (main effect of sex, p=0.005 and p<0.0001, respectively; Table 2). Sex differences in hippocampal volume were observed regardless of diagnosis or APOE genotype.

### 3.6. Diagnosis is associated with Aβ and executive function

Irrespective of sex and APOE genotype, increasing severity of diagnosis was associated with lower CSF Aβ levels in LMCI and probable AD compared to CN individuals (p<0.0001). Aβ levels were similar in CN and EMCI individuals (p=0.1). Increasing severity of diagnosis was associated with lower executive function z-scores (p<0.0001), irrespective of sex and APOE genotype.

## 4. Discussion

In the present study, we found that APOE genotype affects AD neuropathology differently in males and females. CSF tau pathology was disproportionately elevated in female carriers of APOEε4 alleles, whereas CSF Aβ was disproportionately reduced in male carriers of APOEε4 alleles. These findings suggest that APOE genotype leads to greater AD neuropathology in both sexes that depends on the pathology biomarker, with females more likely to show higher CSF tau levels and males to present with lower CSF Aβ levels (suggesting a higher Aβ deposition). Females displayed higher memory scores than males in CN, EMCI, and LMCI groups but not in probable AD, which may contribute to delayed diagnosis in females (Sundermann et al., 2017) and/or to the more severe decline in memory across diagnosis in females compared to males (Buckley et al., 2018; Irvine et al., 2012; Wang et al., 2019). Interestingly, depending on the correction method, females had either smaller or larger hippocampus volume compared to males, regardless of APOE genotype or diagnosis. As expected, increasing severity of diagnosis reduced executive function and CSF Aβ, irrespective of sex and APOE genotype, and APOE genotype affected memory, hippocampal volume, and tau-pathology depending on diagnosis. Previous work has demonstrated sex differences in rates of AD and symptoms of AD (reviewed in Ferretti et al., 2018; Nebel et al., 2018). Our current study suggests that AD biomarkers (CSF tau and Aβ) are expressed differently between the sexes with APOE genotype, suggesting different AD pathology pathways between females and males carriers of APOEε4 alleles. These data may provide clues to better understand the sex differences in AD lifetime risk and progression.

### 4.1 APOE genotype affects tau and Aβ pathology differently between the sexes

We found that levels of tau and p-tau were disproportionately elevated with APOEε4 allele expression in females compared to males. Previous ADNI studies have found a significant interaction between sex and APOE genotype on levels of CSF tau in CN (Damoiseaux et al., 2012; but see Sampedro et al., 2015; Buckley et al., 2019a; Altmann et al., 2014) and aMCI participants (Altmann et al., 2014). A recent meta-analysis found a stronger association between APOEε4 and tau levels in CN females compared to males using ADNI and other datasets (Hohman et al., 2018). Our data extends these data by adding that the interaction between sex and APOE genotype on CSF tau and ptau is seen regardless of diagnosis (in CN, EMCI, LMCI and AD individuals). A recent study using a different cohort found that CSF ptau levels were not significantly higher in female APOEε4 carriers diagnosed with probable AD but a sex difference was found in earlier stages of the disease, i.e., subjective cognitive decline and MCI (Babapour Mofrad et al., 2020). Previous studies indicate that females with at least one APOEε4 allele are at a greater risk for developing AD earlier than are males with this allele (Altmann et al., 2014; Neu et al., 2017), and sex differences in tau and p-tau may be one underlying mechanism by which this occurs but further replication with additional and larger populations are needed.

In contrast to tau pathology, CSF Aβ levels were disproportionately reduced in male APOEε4 carriers compared to females, suggesting a greater Aβ deposition in male carriers of APOEε4 alleles, regardless of diagnosis. To date, studies have not been consistent regarding sex differences in CSF Aβ levels or Aβ deposition based on APOE genotype. For example, somewhat consistent with our findings some studies have found that male APOEε4 carriers have lower CSF Aβ levels in CN individuals (but not in aMCI individuals; Altman et al., 2014), and that males, regardless of APOE status, have higher Aβ deposition (using PET) than females in certain brain regions (i.e., cingulate cortex; Cavedo et al., 2018). However, other studies in cognitively healthy individuals find no sex differences in Aβ deposition (Jack et al., 2015) or that females have higher levels of Aβ deposition (Sundermann et al., 2018), regardless of APOE genotype. Differences between studies may be due to differences between CSF and PET Aβ levels and types of analysis. Levels of CSF tau are hypothesized to increase after CSF Aβ declines and Aβ aggregates and deposits in the brain (Blennow et al., 2015). The association between tau and Aβ is also well characterized, i.e., increasing tau PET levels are associated with increasing Aβ PET deposition (e.g., Maass et al., 2017). Based on this, we would have expected lower CSF Aβ to be associated with higher CSF tau and ptau, which is precisely what we found in females, regardless of APOE genotype, but only in male non-carriers of APOEε4 alleles. Previous work is consistent with this finding, as higher levels of tau PET were more strongly associated with higher Aβ burden in the entorhinal cortex in females compared to males (Buckley et al., 2019b). Taken together, this suggests that the weaker relationship between tau and Aβ in males is due to APOE genotype and the stronger relationship in females between tau and Aβ is seen regardless of APOE genotype and these sex differences may drive inconsistences between studies. However, CSF tau and Aβ were similarly correlated with memory scores in both sexes and APOE genotype. It is possible that depending on APOE genotype, the neurodegeneration pathway (tau or Aβ) or timeline of pathology is different between the sexes, but whichever pathway is present, it is correlated with lower memory scores.

### 4.2 AD diagnosis decreased memory scores to a greater extent in females

As expected, we found that the presence of APOEε4 alleles and probable AD diagnosis were associated with reduced memory and executive function scores consistent with past literature (Buckner, 2004; Ewers et al., 2012; Petersen et al., 2000). However, memory scores in females were more affected with diagnosis such that the female memory advantage was reduced with probable AD, and this is in line with longitudinal analyses showing females decline faster (although this may depend on APOE genotype and Aβ burden; Holland et al., 2013; Buckley et al., 2018). Executive function on the other hand was similarly reduced in females and males with diagnosis. Previous studies found that females have better verbal memory compared to males across diagnoses (CN, Jack et al., 2015; aMCI and probable AD, Sundermann et al., 2018, 2016). Here, we used the ADNI memory score developed by Crane et al. (2012) to detect abnormal memory including language, attention, and logical memory. It is possible that verbal memory may be driving the sex difference favouring females in the present study, however individual cognitive scores will need to be analysed to confirm this. In contrast, Buckley et al. (2018) found no sex differences using a composite cognitive score that includes memory and executive function (Preclinical Alzheimer’s Cognitive Composite score with semantic processing, PACC5) using ADNI and two other cohorts. Altogether, we found that in females CSF tau pathology was increased with APOE genotype and that in CN, EMCI and LMCI females (regardless of APOE genotype) memory and executive function scores were higher compared to males suggesting females have a reserve against brain pathology that delays either the onset of cognitive decline or diagnosis (Sundermann et al., 2017). Females may also use different coping strategies which delay manifestation of clinical symptoms, and in turn females may be less likely to be diagnosed with MCI or AD, as is the case with other conditions such as cardiovascular disease (Norris et al., 2020). Another study found that demographic differences (education and socioeconomic status) can affect sex differences in cognition. Males score better in verbal learning in impoverished and poor health cohorts and this is also associated with better education in males. In contrast, females score better when they have better education even when using education as a covariate (Hogervorst et al., 2012). These results suggest that more detailed neuropsychological analyses are important to consider, along with the use of composite or individual cognitive scores, and that education is a moderating variable in the effects of sex on neuropsychological measures of cognition. The use of education as a moderating variable to influence cognitive memory scores across diagnosis and genotype will be an important factor in future studies. However, once cognitive decline begins, sex differences in memory scores are reduced (current study) and females show higher rates of declines compared to males (Buckley et al., 2018; Holland et al., 2013; Hua et al., 2010) perhaps because the underlying tau pathology is elevated in females.

Females have a higher lifetime risk of AD than males and show greater cognitive decline with AD than males (reviewed by Ferretti et al., 2018; Nebel et al., 2018). The disproportionate effect in females compared to males of (1) APOEε4 genotype on tau-related pathology and (2) AD diagnosis on memory scores supports the idea that females have a higher burden of the disease, although not all studies agree (Jack et al., 2019; reviewed in Nebel et al., 2018). Intriguingly, males are more likely to be diagnosed with aMCI compared to females (Jack et al., 2019), whereas females progress faster from aMCI to AD (Lin et al., 2015). These findings are similar to those in the current study as we found more males than females diagnosed with LMCI but no sex difference in frequency in probable AD. Altogether, our study and previous research suggests that AD females “catch up” to males and sex differences in tau-related pathology found in the current study may be the underlying mechanism for this accelerated transition.

### 4.3 Sex differences in hippocampal volume depended on correction method

We found that the presence of APOEε4 alleles and probable AD diagnosis were associated with reduced corrected hippocampal volume (both methods of correction) consistent with previous literature (Buckner, 2004; Ewers et al., 2012; Jack et al., 2000; Li et al., 2016; Petersen et al., 2000). Curiously, depending on the correction method, females had larger or smaller hippocampal volume compared to males. Females had smaller corrected hippocampal volume using the residuals of the linear regression between hippocampal volume and total intracranial volume (regression method) but larger ratio of hippocampal/intracranial volume compared to males. Similar findings have been shown before as sex differences depend on whether hippocampal volume is corrected for by intracranial volume or total brain volume, and the method of correction (Tan et al., 2016). In their meta-analysis, Tan et al. (2016) conclude that sex differences in hippocampal volume are modest and highly depends on whether or not a correction was applied and the method of correction which is supported by our findings. These results have important implications for understanding sex differences in diseases that influence the integrity of the hippocampus as they lead to very different conclusions regarding hippocampal plasticity.

### 4.4 Limitations

The ADNI data are not representative of the population as it is mostly composed of white and highly educated individuals. As ethnicity (Mayeda et al., 2016; Steenland et al., 2016) and education (Sharp and Gatz, 2011) can affect incidence, prevalence and age of AD onset, our conclusions may not apply to more ethnically and socially diverse populations. Although we controlled for education levels, education in itself is an important contributing factor and may impact sex differences in cognitive decline (Hogervorst et al., 2012). It is also possible that the protective effects of education vary with race/ethnicity (Avila et al., 2021), therefore future studies in more representative populations are needed. In addition, other pathologies in these participants, such as cancer, cardiovascular disease, diabetes, or smoking status may have influenced our measures of AD pathology, cognition, and hippocampal volume (Durazzo et al., 2014; Mezencev and Chernoff, 2020; Santiago and Potashkin, 2021). In this cohort, we did not detect significant interactions between sex, APOE genotype, and diagnosis for any of the endpoints analysed. Future research with large cohorts is required to further test how sex, APOE genotype, and diagnosis interact together as well as how other life history characteristics such as parity may play a role.

## 6. Conclusion

Levels of CSF tau and p-tau were disproportionately affected by APOE genotype in females compared to males, however males had a higher reduction in CSF Aβ levels with APOE genotype compared to females. Our results support the idea that there are sex differences in the manifestation and pathology of AD with APOE genotype. Interestingly, although in this cohort female APOEε4 carriers had elevated tau pathology, females (regardless of APOE genotype) had higher memory scores compared to males in CN and aMCI groups. Therefore, it is possible that females may have a reserve that protects the brain from damage to delay cognitive decline or delay diagnosis but as the disease progresses the advantage in memory is reduced and females show a faster trajectory of cognitive decline.

## Data Availability

Data used in preparation of this article were obtained from the Alzheimer's Disease Neuroimaging Initiative (ADNI) database (adni.loni.usc.edu).

## Acknowledgments

Data collection and sharing for this project was funded by the Alzheimer’s Disease Neuroimaging Initiative (ADNI) (National Institutes of Health Grant U01 AG024904) and DOD ADNI (Department of Defense award number W81XWH-12-2-0012). ADNI is funded by the National Institute on Aging, the National Institute of Biomedical Imaging and Bioengineering, and through generous contributions from the following: AbbVie, Alzheimer’s Association; Alzheimer’s Drug Discovery Foundation; Araclon Biotech; BioClinica, Inc.; Biogen; Bristol-Myers Squibb Company; CereSpir, Inc.; Cogstate; Eisai Inc.; Elan Pharmaceuticals, Inc.; Eli Lilly and Company; EuroImmun; F. Hoffmann-La Roche Ltd and its affiliated company Genentech, Inc.; Fujirebio; GE Healthcare; IXICO Ltd.; Janssen Alzheimer Immunotherapy Research & Development, LLC.; Johnson & Johnson Pharmaceutical Research & Development LLC.; Lumosity; Lundbeck; Merck & Co., Inc.; Meso Scale Diagnostics, LLC.; NeuroRx Research; Neurotrack Technologies; Novartis Pharmaceuticals Corporation; Pfizer Inc.; Piramal Imaging; Servier; Takeda Pharmaceutical Company; and Transition Therapeutics. The Canadian Institutes of Health Research is providing funds to support ADNI clinical sites in Canada. Private sector contributions are facilitated by the Foundation for the National Institutes of Health (www.fnih.org). The grantee organization is the Northern California Institute for Research and Education, and the study is coordinated by the Alzheimer’s Therapeutic Research Institute at the University of Southern California. ADNI data are disseminated by the Laboratory for Neuro Imaging at the University of Southern California. Funding for this study was provided by a Canadian Institutes of Health Research (CHIR) grant to LAMG (PJT-148662). PDG is funded by the Alzheimer’s Association of the USA and Brain Canada with the financial support of Health Canada through the Brain Canada Research Fund (AARF-17-529705). The views expressed herein do not necessarily represent the views of the Minister of Health or the Government of Canada.

## References

Altmann, A., Tian, L., Henderson, V.W., Greicius, M.D., 2014. Sex modifies the APOE-related risk of developing Alzheimer disease. Annals of Neurology 75, 563–573. https://doi.org/10.1002/ana.24135

Alzheimer’s Association, 2017. 2017 Alzheimer’s disease facts and figures. Alzheimer’s & Dementia 13, 325–373. https://doi.org/10.1016/j.jalz.2017.02.001

Ardekani, B.A., Convit, A., Bachman, A.H., 2016. Analysis of the MIRIAD Data Shows Sex Differences in Hippocampal Atrophy Progression. J. Alzheimers Dis. 50, 847–857. https://doi.org/10.3233/JAD-150780

Asperholm, M., Högman, N., Rafi, J., Herlitz, A., 2019. What did you do yesterday? A meta-analysis of sex differences in episodic memory. Psychol Bull 145, 785–821. https://doi.org/10.1037/bul0000197

Avila, J.F., Rentería, M.A., Jones, R.N., Vonk, J.M.J., Turney, I., Sol, K., Seblova, D., Arias, F., Hill-Jarrett, T., Levy, S.-A., Meyer, O., Racine, A.M., Tom, S.E., Melrose, R.J., Deters, K., Medina, L.D., Carrión, C.I., Díaz-Santos, M., Byrd, D.R., Chesebro, A., Colon, J., Igwe, K.C., Maas, B., Brickman, A.M., Schupf, N., Mayeux, R., Manly, J.J., 2021. Education differentially contributes to cognitive reserve across racial/ethnic groups. Alzheimer’s & Dementia 17, 70–80. https://doi.org/10.1002/alz.12176

Babapour Mofrad, R., Tijms, B.M., Scheltens, P., Barkhof, F., van der Flier, W.M., Sikkes, S.A.M., Teunissen, C.E., 2020. Sex differences in CSF biomarkers vary by Alzheimer disease stage and APOE ε4 genotype. Neurology 95, e2378–e2388. https://doi.org/10.1212/WNL.0000000000010629

Benjamini, Y., Hochberg, Y., 1995. Controlling the False Discovery Rate: A Practical and Powerful Approach to Multiple Testing. Journal of the Royal Statistical Society.Series B (Methodological) 57, 289–300.

Blennow, K., Dubois, B., Fagan, A.M., Lewczuk, P., de Leon, M.J., Hampel, H., 2015. Clinical utility of cerebrospinal fluid biomarkers in the diagnosis of early Alzheimer’s disease. Alzheimer’s & Dementia 11, 58–69. https://doi.org/10.1016/j.jalz.2014.02.004

Buckley, R.F., Mormino, E.C., Amariglio, R.E., Properzi, M.J., Rabin, J.S., Lim, Y.Y., Papp, K.V., Jacobs, H.I.L., Burnham, S., Hanseeuw, B.J., Doré, V., Dobson, A., Masters, C.L., Waller, M., Rowe, C.C., Maruff, P., Donohue, M.C., Rentz, D.M., Kirn, D., Hedden, T., Chhatwal, J., Schultz, A.P., Johnson, K.A., Villemagne, V.L., Sperling, R.A., 2018. Sex, amyloid, and APOE ε4 and risk of cognitive decline in preclinical Alzheimer’s disease: Findings from three well-characterized cohorts. Alzheimer’s & Dementia 14, 1193–1203. https://doi.org/10.1016/j.jalz.2018.04.010

Buckley, R.F., Mormino, E.C., Chhatwal, J., Schultz, A.P., Rabin, J.S., Rentz, D.M., Acar, D., Properzi, M.J., Dumurgier, J., Jacobs, H., Gomez-Isla, T., Johnson, K.A., Sperling, R.A., Hanseeuw, B.J., 2019a. Associations between baseline amyloid, sex, and APOE on subsequent tau accumulation in cerebrospinal fluid. Neurobiology of Aging 78, 178–185. https://doi.org/10.1016/j.neurobiolaging.2019.02.019

Buckley, R.F., Mormino, E.C., Rabin, J.S., Hohman, T.J., Landau, S., Hanseeuw, B.J., Jacobs, H.I.L., Papp, K.V., Amariglio, R.E., Properzi, M.J., Schultz, A.P., Kirn, D., Scott, M.R., Hedden, T., Farrell, M., Price, J., Chhatwal, J., Rentz, D.M., Villemagne, V.L., Johnson, K.A., Sperling, R.A., 2019b. Sex Differences in the Association of Global Amyloid and Regional Tau Deposition Measured by Positron Emission Tomography in Clinically Normal Older Adults. JAMA Neurol 76, 542–551. https://doi.org/10.1001/jamaneurol.2018.4693

Buckner, R.L., 2004. Memory and Executive Function in Aging and AD: Multiple Factors that Cause Decline and Reserve Factors that Compensate. Neuron 44, 195–208. https://doi.org/10.1016/j.neuron.2004.09.006

Cavedo, E., Chiesa, P.A., Houot, M., Ferretti, M.T., Grothe, M.J., Teipel, S.J., Lista, S., Habert, M.-O., Potier, M.-C., Dubois, B., Hampel, H., Bakardjian, H., Benali, H., Bertin, H., Bonheur, J., Boukadida, L., Boukerrou, N., Cavedo, E., Chiesa, P., Colliot, O., Dubois, B., Dubois, M., Epelbaum, S., Gagliardi, G., Genthon, R., Habert, M.-O., Hampel, H., Houot, M., Kas, A., Lamari, F., Levy, M., Lista, S., Metzinger, C., Mochel, F., Nyasse, F., Poisson, C., Potier, M.-C., Revillon, M., Santos, A., Andrade, K.S., Sole, M., Surtee, M., de Schotten, M.T., Vergallo, A., Younsi, N., Aguilar, L.F., Babiloni, C., Baldacci, F., Benda, N., Black, K.L., Bokde, A.L.W., Bonuccelli, U., Broich, K., Bun, R.S., Cacciola, F., Castrillo, J., Cavedo, E., Ceravolo, R., Chiesa, P.A., Colliot, O., Coman, C.-M., Corvol, J.-C., Cuello, A.C., Cummings, J.L., Depypere, H., Dubois, B., Duggento, A., Durrleman, S., Escott-Price, V., Federoff, H., Ferretti, M.T., Fiandaca, M., Frank, R.A., Garaci, F., Genthon, R., George, N., Giorgi, F.S., Graziani, M., Haberkamp, M., Habert, M.-O., Hampel, H., Herholz, K., Karran, E., Kim, S.H., Koronyo, Y., Koronyo-Hamaoui, M., Lamari, F., Langevin, T., Lehéricy, S., Lista, S., Lorenceau, J., Mapstone, M., Neri, C., Nisticò, R., Nyasse-Messene, F., O’bryant, S.E., Perry, G., Ritchie, C., Rojkova, K., Rossi, S., Saidi, A., Santarnecchi, E., Schneider, L.S., Sporns, O., Toschi, N., Verdooner, S.R., Vergallo, A., Villain, N., Welikovitch, L.A., Woodcock, J., Younesi, E., 2018. Sex differences in functional and molecular neuroimaging biomarkers of Alzheimer’s disease in cognitively normal older adults with subjective memory complaints. Alzheimer’s & Dementia 14, 1204–1215. https://doi.org/10.1016/j.jalz.2018.05.014

Crane, P.K., Carle, A., Gibbons, L.E., Insel, P., Mackin, R.S., Gross, A., Jones, R.N., Mukherjee, S., Curtis, S.M., Harvey, D., Weiner, M., Mungas, D., Alzheimer’s Disease Neuroimaging Initiative, 2012. Development and assessment of a composite score for memory in the Alzheimer’s Disease Neuroimaging Initiative (ADNI). Brain Imaging Behav 6, 502–516. https://doi.org/10.1007/s11682-012-9186-z

Damoiseaux, J.S., Seeley, W.W., Zhou, J., Shirer, W.R., Coppola, G., Karydas, A., Rosen, H.J., Miller, B.L., Kramer, J.H., Greicius, M.D., Alzheimer’s Disease Neuroimaging Initiative, 2012. Gender modulates the APOE ε4 effect in healthy older adults: convergent evidence from functional brain connectivity and spinal fluid tau levels. J. Neurosci. 32, 8254–8262. https://doi.org/10.1523/JNEUROSCI.0305-12.2012

Durazzo, T.C., Mattsson, N., Weiner, M.W., 2014. Smoking and increased Alzheimer’s disease risk: A review of potential mechanisms. Alzheimers Dement 10, S122–S145. https://doi.org/10.1016/j.jalz.2014.04.009

Ewers, M., Walsh, C., Trojanowski, J.Q., Shaw, L.M., Petersen, R.C., Jack, C.R., Feldman, H.H., Bokde, A.L.W., Alexander, G.E., Scheltens, P., Vellas, B., Dubois, B., Weiner, M., Hampel, H., 2012. Prediction of conversion from mild cognitive impairment to Alzheimer’s disease dementia based upon biomarkers and neuropsychological test performance. Neurobiology of Aging 33, 1203-1214.e2. https://doi.org/10.1016/j.neurobiolaging.2010.10.019

Ferretti, M.T., Iulita, M.F., Cavedo, E., Chiesa, P.A., Schumacher Dimech, A., Santuccione Chadha, A., Baracchi, F., Girouard, H., Misoch, S., Giacobini, E., Depypere, H., Hampel, H., 2018. Sex differences in Alzheimer disease — the gateway to precision medicine. Nature Reviews Neurology 14, 457–469. https://doi.org/10.1038/s41582-018-0032-9

Gaillard, A., Fehring, D.J., Rossell, S.L., 2020. A systematic review and meta-analysis of behavioural sex differences in executive control. European Journal of Neuroscience. https://doi.org/10.1111/ejn.14946

Gibbons, L.E., Carle, A.C., Mackin, R.S., Harvey, D., Mukherjee, S., Insel, P., Curtis, S.M., Mungas, D., Crane, P.K., Alzheimer’s Disease Neuroimaging Initiative, 2012. A composite score for executive functioning, validated in Alzheimer’s Disease Neuroimaging Initiative (ADNI) participants with baseline mild cognitive impairment. Brain Imaging Behav 6, 517–527. https://doi.org/10.1007/s11682-012-9176-1

Hogervorst, E., Rahardjo, T.B., Jolles, J., Brayne, C., Henderson, V.W., 2012. Gender differences in verbal learning in older participants. Aging Health 8, 493–507. https://doi.org/10.2217/ahe.12.56

Hohman, T.J., Dumitrescu, L., Barnes, L.L., Thambisetty, M., Beecham, G., Kunkle, B., Gifford, K.A., Bush, W.S., Chibnik, L.B., Mukherjee, S., Jager, P.L.D., Kukull, W., Crane, P.K., Resnick, S.M., Keene, C.D., Montine, T.J., Schellenberg, G.D., Haines, J.L., Zetterberg, H., Blennow, K., Larson, E.B., Johnson, S.C., Albert, M., Bennett, D.A., Schneider, J.A., Jefferson, A.L., 2018. Sex-Specific Association of Apolipoprotein E With Cerebrospinal Fluid Levels of Tau. JAMA Neurol 75, 989–998. https://doi.org/10.1001/jamaneurol.2018.0821

Holland, D., Desikan, R.S., Dale, A.M., McEvoy, L.K., Alzheimer’s Disease Neuroimaging Initiative, 2013. Higher rates of decline for women and apolipoprotein E epsilon4 carriers. AJNR Am J Neuroradiol 34, 2287–2293. https://doi.org/10.3174/ajnr.A3601

Hua, X., Hibar, D.P., Lee, S., Toga, A.W., Jack, C.R., Weiner, M.W., Thompson, P.M., 2010. Sex and age differences in atrophic rates: an ADNI study with n=1368 MRI scans. Neurobiology of Aging 31, 1463–1480. https://doi.org/10.1016/j.neurobiolaging.2010.04.033

Irvine, K., Laws, K.R., Gale, T.M., Kondel, T.K., 2012. Greater cognitive deterioration in women than men with Alzheimer’s disease: a meta analysis. J Clin Exp Neuropsychol 34, 989–998. https://doi.org/10.1080/13803395.2012.712676

Jack, C.R., Dickson, D.W., Parisi, J.E., Xu, Y.C., Cha, R.H., O’Brien, P.C., Edland, S.D., Smith, G.E., Boeve, B.F., Tangalos, E.G., Kokmen, E., Petersen, R.C., 2002. Antemortem MRI Findings Correlate with Hippocampal Neuropathology in Typical Aging and Dementia. Neurology 58, 750–757.

Jack, C.R., Knopman, D.S., Weigand, S.D., Wiste, H.J., Vemuri, P., Lowe, V., Kantarci, K., Gunter, J.L., Senjem, M.L., Ivnik, R.J., Roberts, R.O., Rocca, W.A., Boeve, B.F., Petersen, R.C., 2012. An operational approach to National Institute on Aging–Alzheimer’s Association criteria for preclinical Alzheimer disease. Annals of Neurology 71, 765–775. https://doi.org/10.1002/ana.22628

Jack, C.R., Petersen, R.C., Xu, Y., O’Brien, P.C., Smith, G.E., Ivnik, R.J., Boeve, B.F., Tangalos, E.G., Kokmen, E., 2000. Rates of hippocampal atrophy correlate with change in clinical status in aging and AD. Neurology 55, 484–489. https://doi.org/10.1212/wnl.55.4.484

Jack, C.R., Therneau, T.M., Weigand, S.D., Wiste, H.J., Knopman, D.S., Vemuri, P., Lowe, V.J., Mielke, M.M., Roberts, R.O., Machulda, M.M., Graff-Radford, J., Jones, D.T., Schwarz, C.G., Gunter, J.L., Senjem, M.L., Rocca, W.A., Petersen, R.C., 2019. Prevalence of Biologically vs Clinically Defined Alzheimer Spectrum Entities Using the National Institute on Aging–Alzheimer’s Association Research Framework. JAMA Neurol. https://doi.org/10.1001/jamaneurol.2019.1971

Jack, C.R., Wiste, H.J., Weigand, S.D., Knopman, D.S., Vemuri, P., Mielke, M.M., Lowe, V., Senjem, M.L., Gunter, J.L., Machulda, M.M., Gregg, B.E., Pankratz, V.S., Rocca, W.A., Petersen, R.C., 2015. Age, Sex, and APOE ε4 Effects on Memory, Brain Structure, and β-Amyloid Across the Adult Life Span. JAMA Neurol 72, 511–519. https://doi.org/10.1001/jamaneurol.2014.4821

Kidron, D., Black, S.E., Stanchev, P., Buck, B., Szalai, J.P., Parker, J., Szekely, C., Bronskill, M.J., 1997. Quantitative MR volumetry in Alzheimer’s disease: Topographic markers and the effects of sex and education. Neurology 49, 1504–1512. https://doi.org/10.1212/WNL.49.6.1504

Li, B., Shi, J., Gutman, B.A., Baxter, L.C., Thompson, P.M., Caselli, R.J., Wang, Y., 2016. Influence of APOE Genotype on Hippocampal Atrophy over Time - An N=1925 Surface-Based ADNI Study. PLoS One 11. https://doi.org/10.1371/journal.pone.0152901

Lin, K.A., Choudhury, K.R., Rathakrishnan, B.G., Marks, D.M., Petrella, J.R., Doraiswamy, P.M., 2015. Marked gender differences in progression of mild cognitive impairment over 8 years. Alzheimer’s & Dementia: Translational Research & Clinical Interventions 1, 103–110. https://doi.org/10.1016/j.trci.2015.07.001

Liu, M., Paranjpe, M.D., Zhou, X., Duy, P.Q., Goyal, M.S., Benzinger, T.L.S., Lu, J., Wang, R., Zhou, Y., 2019. Sex modulates the ApoE ε4 effect on brain tau deposition measured by 18F-AV-1451 PET in individuals with mild cognitive impairment. Theranostics 9, 4959–4970. https://doi.org/10.7150/thno.35366

Lotze, M., Domin, M., Gerlach, F.H., Gaser, C., Lueders, E., Schmidt, C.O., Neumann, N., 2019. Novel findings from 2,838 Adult Brains on Sex Differences in Gray Matter Brain Volume. Scientific Reports 9, 1671. https://doi.org/10.1038/s41598-018-38239-2

Maass, A., Landau, S., Baker, S.L., Horng, A., Lockhart, S.N., La Joie, R., Rabinovici, G.D., Jagust, W.J., 2017. Comparison of multiple tau-PET measures as biomarkers in aging and Alzheimer’s disease. NeuroImage 157, 448–463. https://doi.org/10.1016/j.neuroimage.2017.05.058

Mayeda, E.R., Glymour, M.M., Quesenberry, C.P., Whitmer, R.A., 2016. Inequalities in dementia incidence between six racial and ethnic groups over 14 years. Alzheimer’s & Dementia 12, 216–224. https://doi.org/10.1016/j.jalz.2015.12.007

Mezencev, R., Chernoff, Y.O., 2020. Risk of Alzheimer’s Disease in Cancer Patients: Analysis of Mortality Data from the US SEER Population-Based Registries. Cancers (Basel) 12. https://doi.org/10.3390/cancers12040796

Mormino, E.C., Betensky, R.A., Hedden, T., Schultz, A.P., Amariglio, R.E., Rentz, D.M., Johnson, K.A., Sperling, R.A., 2014. Synergistic Effect of β-Amyloid and Neurodegeneration on Cognitive Decline in Clinically Normal Individuals. JAMA Neurol 71, 1379–1385. https://doi.org/10.1001/jamaneurol.2014.2031

Nebel, R.A., Aggarwal, N.T., Barnes, L.L., Gallagher, A., Goldstein, J.M., Kantarci, K., Mallampalli, M.P., Mormino, E.C., Scott, L., Yu, W.H., Maki, P.M., Mielke, M.M., 2018. Understanding the impact of sex and gender in Alzheimer’s disease: A call to action. Alzheimer’s & Dementia 14, 1171–1183. https://doi.org/10.1016/j.jalz.2018.04.008

Neu, S.C., Pa, J., Kukull, W., Beekly, D., Kuzma, A., Gangadharan, P., Wang, L.-S., Romero, K., Arneric, S.P., Redolfi, A., Orlandi, D., Frisoni, G.B., Au, R., Devine, S., Auerbach, S., Espinosa, A., Boada, M., Ruiz, A., Johnson, S.C., Koscik, R., Wang, J.-J., Hsu, W.-C., Chen, Y.-L., Toga, A.W., 2017. Apolipoprotein E Genotype and Sex Risk Factors for Alzheimer Disease: A Meta-analysis. JAMA Neurol. https://doi.org/10.1001/jamaneurol.2017.2188

Norris, C.M., Yip, C.Y.Y., Nerenberg, K.A., Clavel, M.-A., Pacheco, C., Foulds, H.J.A., Hardy, M., Gonsalves, C.A., Jaffer, S., Parry, M., Colella, T.J.F., Dhukai, A., Grewal, J., Price, J.A.D., Levinsson, A.L.E., Hart, D., Harvey, P.J., Van Spall, H.G.C., Sarfi, H., Sedlak, T.L., Ahmed, S.B., Baer, C., Coutinho, T., Edwards, J.D., Green, C.R., Kirkham, A.A., Srivaratharajah, K., Dumanski, S., Keeping-Burke, L., Lappa, N., Reid, R.D., Robert, H., Smith, G., Martin-Rhee, M., Mulvagh, S.L., 2020. State of the Science in Women’s Cardiovascular Disease: A Canadian Perspective on the Influence of Sex and Gender. J Am Heart Assoc 9, e015634. https://doi.org/10.1161/JAHA.119.015634

Petersen, R.C., Aisen, P.S., Beckett, L.A., Donohue, M.C., Gamst, A.C., Harvey, D.J., Jack, C.R., Jagust, W.J., Shaw, L.M., Toga, A.W., Trojanowski, J.Q., Weiner, M.W., 2010. Alzheimer’s Disease Neuroimaging Initiative (ADNI). Neurology 74, 201–209. https://doi.org/10.1212/WNL.0b013e3181cb3e25

Petersen, R.C., Jack, C.R., Xu, Y.C., Waring, S.C., O’Brien, P.C., Smith, G.E., Ivnik, R.J., Tangalos, E.G., Boeve, B.F., Kokmen, E., 2000. Memory and MRI-based hippocampal volumes in aging and AD. Neurology 54, 581–587. https://doi.org/10.1212/wnl.54.3.581

R Core Team, 2018. A language and environment for statistical computing. R Foundation for Statistical Computing, Vienna, Austria. URL https://www.R-project.org/.

Riedel, B.C., Thompson, P.M., Brinton, R.D., 2016. Age, APOE and sex: Triad of risk of Alzheimer’s disease. The Journal of Steroid Biochemistry and Molecular Biology 160, 134–147. https://doi.org/10.1016/j.jsbmb.2016.03.012

Sampedro, F., Vilaplana, E., de Leon, M.J., Alcolea, D., Pegueroles, J., Montal, V., Carmona-Iragui, M., Sala, I., Sánchez-Saudinos, M.-B., Antón-Aguirre, S., Morenas-Rodríguez, E., Camacho, V., Falcón, C., Pavía, J., Ros, D., Clarimón, J., Blesa, R., Lleó, A., Fortea, J., Alzheimer’s Disease Neuroimaging Initiative, 2015. APOE-by-sex interactions on brain structure and metabolism in healthy elderly controls. Oncotarget 6, 26663–26674. https://doi.org/10.18632/oncotarget.5185

Santiago, J.A., Potashkin, J.A., 2021. The Impact of Disease Comorbidities in Alzheimer’s Disease. Front. Aging Neurosci. 13. https://doi.org/10.3389/fnagi.2021.631770

Sharp, E.S., Gatz, M., 2011. The Relationship between Education and Dementia An Updated Systematic Review. Alzheimer Dis Assoc Disord 25, 289–304. https://doi.org/10.1097/WAD.0b013e318211c83c

Shaw, L.M., Vanderstichele, H., Knapik-Czajka, M., Clark, C.M., Aisen, P.S., Petersen, R.C., Blennow, K., Soares, H., Simon, A., Lewczuk, P., Dean, R., Siemers, E., Potter, W., Lee, V.M.-Y., Trojanowski, J.Q., 2009. Cerebrospinal Fluid Biomarker Signature in Alzheimer’s Disease Neuroimaging Initiative Subjects. Ann Neurol 65, 403–413. https://doi.org/10.1002/ana.21610

Sohn, D., Shpanskaya, K., Lucas, J.E., Petrella, J.R., Saykin, A.J., Tanzi, R.E., Samatova, N.F., Doraiswamy, P.M., 2018. Sex Differences in Cognitive Decline in Subjects with High Likelihood of Mild Cognitive Impairment due to Alzheimer’s disease. Scientific Reports 8, 7490. https://doi.org/10.1038/s41598-018-25377-w

Steenland, K., Goldstein, F.C., Levey, A., Wharton, W., 2016. A Meta-Analysis of Alzheimer’s Disease Incidence and Prevalence Comparing African-Americans and Caucasians. J. Alzheimers Dis. 50, 71–76. https://doi.org/10.3233/JAD-150778

Sundermann, E.E., Biegon, A., Rubin, L.H., Lipton, R.B., Landau, S., Maki, P.M., Alzheimer’s Disease Neuroimaging Initiative, 2017. Does the Female Advantage in Verbal Memory Contribute to Underestimating Alzheimer’s Disease Pathology in Women versus Men? J. Alzheimers Dis. 56, 947–957. https://doi.org/10.3233/JAD-160716

Sundermann, E.E., Biegon, A., Rubin, L.H., Lipton, R.B., Mowrey, W., Landau, S., Maki, P.M., 2016. Better verbal memory in women than men in MCI despite similar levels of hippocampal atrophy. Neurology 86, 1368–1376. https://doi.org/10.1212/WNL.0000000000002570

Sundermann, E.E., Tran, M., Maki, P.M., Bondi, M.W., 2018. Sex differences in the association between apolipoprotein E ε4 allele and Alzheimer’s disease markers. Alzheimer’s & Dementia: Diagnosis, Assessment & Disease Monitoring 10, 438–447. https://doi.org/10.1016/j.dadm.2018.06.004

Tan, A., Ma, W., Vira, A., Marwha, D., Eliot, L., 2016. The human hippocampus is not sexually-dimorphic: Meta-analysis of structural MRI volumes. NeuroImage 124, 350–366. https://doi.org/10.1016/j.neuroimage.2015.08.050

Wang, X., Zhou, W., Ye, T., Lin, X., Zhang, J., Initiative, for A.D.N., 2019. Sex Difference in the Association of APOE4 with Memory Decline in Mild Cognitive Impairment. Journal of Alzheimer’s Disease 69, 1161–1169. https://doi.org/10.3233/JAD-181234

Yagi, S., Galea, L.A.M., 2019. Sex differences in hippocampal cognition and neurogenesis. Neuropsychopharmacology 44, 200–213. https://doi.org/10.1038/s41386-018-0208-4

